# IL-21/23 axis modulates inflammatory cytokines and RANKL expression in RA CD4^+^ T cells via p-Akt signaling

**DOI:** 10.1101/2023.03.29.23287939

**Authors:** Gargee Bhattacharya, Soumya Sengupta, Rohila Jha, Shubham K Shaw, Gajendra M Jogdand, Prakash K Barik, Prasanta Padhan, Jyoti R Parida, Satish Devadas

## Abstract

The role of CD4^+^ T cells specifically, Th17 has been well documented in RA pathogenesis. Here we focus on the critical role of cytokines IL-21 and IL-23 in facilitating the aberrant status of RA Th17-like cells and report their significant contribution(s) in modulating the expression of inflammatory cytokines and RANKL. Neutralizing IL-21 or IL- 23 (p19 and p40) or both, resulted in downregulation of the cytokines, TNF-α, IFN-γ and IL- 17 and RANKL expression in RA CD4^+^ T cells. Our *ex vivo* human Th17 studies also validated the above findings and we hypothesize a common pathway responsible for regulation of inflammatory cytokines and RANKL expression. Subsequent dissection of the signalling pathway found p-Akt1 as the key phosphoprotein downstream of both IL-21 and IL-23, capable of augmenting inflammatory cytokines and RANKL production. Altogether, these findings identify IL-21/23 axis in RA CD4^+^ T cells as a key regulator dictating two critical processes i.e. exaggerated inflammation and higher osteoclastogenesis and provide critical targets in their downstream signalling for therapeutic approaches.

## Introduction

Rheumatoid arthritis (RA) is a chronic autoimmune disorder, essentially characterized by synovial inflammation, bone and cartilage destruction and autoantibody generation (Firestein 2003, Firestein and McInnes 2017). The above immune pathologies are an extensive manifestation of the inflammatory lesions formed as a consequence of immune cell infiltration and their autoimmune response against joint specific antigens, eventually leading to irreversible tissue damage (McInnes and Schett 2011, Jang, Kwon et al. 2022). Although, RA microenvironment is primarily constituted by an array of immune cells, the disease pathology is precipitated by CD4^+^ T cells and more specifically, is Th17-driven where it has been shown that most of the Th17 secreted cytokines, including IL-17, IL-22, TNF-α, IFN-γ are inflammatory in nature (Firestein 2003, Mellado, Martinez-Munoz et al. 2015, Yasuda, Takeuchi et al. 2019). Not surprisingly, hyper secretion of these cytokines, along with uncontrolled proliferation and phenotype aberrancies attribute to the pathogenic Th17 (pTh17) population, distinct from its relevant counterpart in physiology (Lee, Awasthi et al. 2012, Lee, Hall et al. 2020, Wu and Wan 2020). Interestingly, previous reports suggest that these aberrancies are not only limited to the infiltrated population of pathogenic Th17s, but are also found in the circulating ones (Penatti, Facciotti et al. 2017, Maeda, Osaga et al. 2019). In addition, these cells have disrepute for stimulating other immune cells such as macrophages, osteoblasts, chondrocytes and fibroblasts for exacerbated secretion of inflammatory cytokines, thus maintaining an inflammatory nexus in the RA joint (Yang, Qian et al. 2019). While, correlation of IL-17/23 axis with pTh17 population’s hyper inflammatory status has been well documented in RA, implications of other cytokines present in the synovial *milieu* is still unclear.

Along with inflammation, pathogenic Th17 cells are actively involved in bone erosion and cartilage destruction in RA (Kim, Kim et al. 2015, Komatsu and Takayanagi 2018). While these cells are known to take an indirect approach of tissue destruction by stimulating synovial fibroblasts via IL-17, TNF-α, IL-6, etc. they are also directly responsible for higher osteoclastogenesis and in turn, aggravated and inflamed bone erosion (Kotake, Udagawa et al. 1999). This process of osteoclastogenesis is critically regulated by a member of TNF subfamily i.e. Receptor Activator of Nuclear factor κB (RANK) and its ligand RANKL, where binding of RANKL to RANK activates a signalling cascade, triggering osteoclast differentiation and proliferation (Sato, Suematsu et al. 2006). T cells are known secretors of RANKL without bearing RANK receptor suggesting paracrine functions for RANKL including bone and dendritic cell (DC) maturation, breast development, etc. (Akiyama, Shinzawa et al. 2012). However, significantly higher than normal RANKL secretion in RA contributes to higher osteoclastogenic activity in RA and could have both T and osteoblast origins. Although previous studies have shown direct or indirect association of Th17 cells and higher osteoclastogenic activity in RA synovium (Kotake, Udagawa et al. 1999, Sato, Suematsu et al. 2006), most of them basically emphasize on RANKL-RANK association, its downstream signalling pathways and the consequent deleterious implications. Insights into the cellular events preceding RANKL upregulation in RA CD4^+^ T cells or Th17 cells remain unclear and establishing this cellular pathway, and downstream signalling is critical for exploiting the cellular or molecular targets contributing to increased bone erosion in RA with respect to pTh17s.

Human Th17 cells are driven by the combination of cytokines IL-6, IL-1β, IL-21, IL- 23 and TGF-β with TCR activation. In normal physiology, each of these cytokines plays a distinct role in facilitating Th17 polarisation where TCR activation, TGF-β and IL-6 initiate Th17 differentiation. Post ROR-γ synthesis, IL-21 is required for sustaining Th17 and IL-17 production and studies suggest, IL-23 downstream of IL-21 stabilizing Th17’s master transcription factor, ROR-γ apart from other factors (Patel and Kuchroo 2015, Bhaumik and Basu 2017, Stadhouders, Lubberts et al. 2018). Not surprisingly, IL-21 and IL-23 have been shown to have multifaceted roles in activation and expansion of the pathogenic Th17 population in RA (Langrish, Chen et al. 2005, Korn, Bettelli et al. 2007, Niu, He et al. 2010, Lubberts 2015, Dinesh and Rasool 2018, Schinocca, Rizzo et al. 2021). Based on this, we essentially addressed the question of plausible modulators regulating two key pillars of RA pathogenesis i.e. inflammation and bone degradation and accordingly examined the IL-21/23 axis and its downstream signalling pathway in RA CD4^+^ T cell population. Altogether, this study is directed towards characterizing an aberrant CD4^+^ T cell with respect to its inflammatory response and osteoclastogenic capacity and subsequently, dissecting the IL- 21/23 axis and its downstream pathway contributing to modulation of inflammatory cytokines and RANKL on these cells.

## Materials and Methods

### Clinical characteristics of RA patients, demography

A total of 67 patients with confirmed clinical RA were recruited from the Out Patient Department of Odisha Arthritis and Rheumatology Centre (OARC) and Kalinga Institute of Medical Sciences (KIMS), based on the 2010 ACR-EULAR classification. Clinical parameters and the medications taken by these patients are tabulated (**Table 1**). In addition, 25 healthy volunteers were enrolled for the study, primarily devoid of any chronic disorders, allergies or infection. Written informed consent was taken from all the subjects of this study prior to sample collection. The study was approved by the Institutional Human Ethics Board, Institute of Life Sciences.

**Table 1.** Clinical parameters

|  | **Healthy Controls**<br>**(n=25)** | **RA Patients**<br>**(n= 67)** |
| --- | --- | --- |
| **Age,years** | 30.9±2.5 | 47.4±11.1 |
| **Female/male, no.s** | 21/4 | 62/5 |
| **Disease duration, years** | N.A | 7.5±3.5 |
| **Rheumatoid factor-**<br>**positive, n (%)** | N.D | 71.6 |
| **Anti-CCP positive, n (%)** | N.D | 55.2 |
| **CRP, mg/dl** | N.D | 49.7±44.1 |
| **ESR, mm/hour** | N.D | 62.9±33.2 |
| **DAS28** | N.A | 5.20±0.9 |
| **Previous medications, n**<br>**(%)** |  |  |
| **Corticosteroids** | N.A | 38(67) |
| **DMARDS** | N.A |  |
| **Methotrexate** |  | 62(67) |
| **HCQS** |  | 49(67) |
| **Leflunomide** |  | 24(67) |
| **NSAIDS** | N.A | 6(67) |

### PBMC and SFMC isolation

Blood and synovial fluid were drawn from RA patients with their consent detailing the study. For PBMC isolation, 5ml of blood was drawn from RA patients and healthy volunteers and PBMCs were isolated by density gradient centrifugation using Histopaque-1077 (Sigma, Burlington, MA, USA) based on manufacturer’s protocol. For isolation of SFMCs (synovial fluid mononuclear cells), synovial fluid was primarily treated with 300µg/ml Hyaluronidase (Sigma, Burlington, MA, USA) for 20 minutes and then layered on Histopaque-1077 for density gradient centrifugation. The isolated PBMCs and SFMCs were counted and cryopreserved at a density of 10 million cells per ml of storage media in liquid Nitrogen for later studies.

### Plasma cytokine detection assay

Neat plasma derived from RA patients and healthy controls was stored in -80^0^C until assayed for cytokines. The samples were run in duplicates to measure 20 T cell cytokines using human Milliplex map cytokine assay kit (Millipore, Billerica, MA, USA) according to manufacturer’s protocol. The samples were acquired in a Bio-Plex 200 system (Bio-Rad, Hercules, CA) and cytokine concentrations were calculated using Bio-Plex manager software with a five-parameter (5PL) curve-fitting algorithm applied for standard curve calculation.

### *Ex vivo* differentiation of human CD4^+^ T cells to Th17 cells

CD4^+^ T cell population was derived from PBMCs by negative selection using Dynabeads Human CD4 T cell kit (Invitrogen, Waltham, MA, USA), according to manufacturer’s protocol. Cell purity was ascertained to be ∼90% as analyzed by flow cytometer and the isolated cells were cultured in RPMI 1640 (PAN-Biotech, Aidenbach, Bavaria, Germany), supplemented with 10% fetal bovine serum, Australian origin (PAN-Biotech, Aidenbach, Bavaria, Germany), 100 U/ml penicillin (Sigma, Burlington, MA, USA), 100 μg/ml streptomycin (Sigma, Burlington, MA, USA) and 50 mM 2β- mercaptoethanol (Sigma, Burlington, MA, USA). For Th17 differentiation, cells were plated on pre-coated αCD3 plates at 1 million per ml density. αCD28 was added in the soluble form (2 μg/ml) along with neutralizing antibodies, αIFN-γ (10 μg/ml), αIL-4 (10 μg/ml), and cytokines IL-6 (25 ng/ml), IL-21 (25 ng/ml), IL-23 (25 ng/ml), IL-1β (15 ng/ml), TGF-β (5ng/ml). The culture was activated for 10 days, followed by washing with RPMI 1640 and used for subsequent experiments.

### Surface markers and intracellular staining of cytokines and transcription factors

For determination of T cell specific surface proteins, secreted cytokines and transcription factors, the cells were activated with PMA (50 ng/ml) (Sigma, Burlington, MA, USA) and Ionomycin (1 μg/ml) (Sigma, Burlington, MA, USA) for 6 hours with Brefeldin A (10 μg/ml) (Sigma, Burlington, MA, USA) being added in the last 4 hours of stimulation. Stimulated cells were stained with dead cell discrimination dye for 20 minutes on ice and then washed. For the analysis of surface proteins, the cells were stained with marker specific antibodies whereas the cells were fixed with Cytofix/Cytoperm Fixation/Permeabilization Solution Kit (BD Biosciences, San Jose, CA, USA) for cytokine staining, and FOXP3 staining buffer set (eBioscience, San Diego, CA, USA), for staining transcription factors. The fixed cells were stained with specific fluorochrome-labelled antibodies for 30 minutes, washed and acquired in BD LSR Fortessa.

### Cytokine block

In brief, PBMCs derived from RA patients were seeded in flat-bottomed plates at 1×10^6^ cells/ml density with neutralization antibodies for the cytokines, IL-21(eBioscience, San Diego, CA, USA), IL-23 p19 (eBioscience, San Diego, CA, USA) and or IL-23 p40 (eBioscience, San Diego, CA, USA) for 30 minutes at room temperature. These cells were then activated with PMA and Ionomycin for subsequent experiments where the cells were either stained for inflammatory cytokines and/or RANKL expression.

### Phospho-Akt1 staining for flow cytometry

For phospho-Akt1 expression, cells were stimulated with αCD3 for 24 hours at room temperature. These cells were then permeabilised with 80% Methanol on ice, washed and fixed with Cytofix/Cytoperm Fixation/Permeabilization Solution Kit (BD Biosciences, San Jose, CA, USA) for 20 minutes at room temperature. Cells were stained with specific antibodies for phospho-Akt1 and kept at room temperature for 30 minutes, washed and acquired in BD LSR Fortessa.

### Phospho-Akt1 inhibition

In brief, CD4^+^ T cells derived from RA PBMCs were stimulated with pre-coated αCD3/28 for 24 hours at room temperature in the presence of phospho-Akt1/2 kinase inhibitor (Sigma, Burlington, MA, USA). These cells were stained for RANKL and inflammatory cytokine expression using the protocol mentioned previously.

### Confocal microscopy

For surface staining of membrane RANKL (mRANKL), CD4^+^ T cells were isolated from PBMCs by using Dynabeads Human CD4 T cell kit (Invitrogen, MA, USA), according to manufacturer’s protocol. Surface staining protocol for CD4 and RANKL was followed as mentioned above. Finally, the cells were stained with Hoechst staining solution (Sigma, Burlington, MA, USA) for 20 minutes and were smeared on pre-coated Poly-L-Lysine slides (Sigma, Burlington, MA, USA). For nuclear translocation studies, isolated CD4^+^ T cells were stimulated with αCD3/28 for 24 hours in the presence or absence of Akt1/2 kinase inhibitor and stained for pAkt1 expression using FOXP3 staining buffer set (eBioscience, San Diego, CA, USA). TCS SP5 Leica confocal microscope was used to visualise surface staining using 488nm and 640nm laser.

### Statistics

All flow cytometric analysis was done in FlowJo software V10.8 (BD Biosciences). Statistical analysis was performed using the GraphPad Prism software, version 9.0.1. Data is presented as Mean ± Standard Error of Mean (SEM). Mann-Whitney U test was used to compare statistics between HC and RA patients including cytokine profile from multiplexing. Paired t test was used to compare between the PMA/Ionomycin stimulated and αIL-21 or αIL- 23 or αIL-21-23 treated cells from same donors. Paired t-test was also used to compare between αCD3/28 stimulated cells and αIL-21 or αIL-23 or αIL-21+αIL-23 treated cells from the same donors. p values less than 0.05 were considered significant (*p < 0.05, **p < 0.01, ***p< 0.001, ****p< 0.0001).

## Results

### Inflammatory cytokine *milieu* defines and dictates Rheumatoid Arthritis

Although RA is essentially characterised by synovitis, it also corresponds to extra-articular manifestations, thus becoming systemic. Accordingly, we set our primary objective to examine the plasma of RA patients with respect to its cytokine profile and compare it to that of the healthy volunteers. Amongst the 20 cytokines analyzed, most of the inflammatory cytokines were found to be significantly elevated in RA (Figure 1A), indicating covert systemic inflammation. Along with this, we also found an increase in the driving cytokines crucial for maintaining RA pathogenesis, such as IL-6, IL-1β and IL-21 (Figure 1B). On the contrary, anti-inflammatory cytokines did not follow a specific trend, where certain cytokines such as IL-13 and IL-10 showed a significant difference whereas IL-4 and IL-5 did not, indicating a skewed or inadequate anti-inflammatory response incapable of containing the inflammation (Figure 1C). In addition, we found elevated levels of certain pleiotropic cytokines responsible for maintaining RA pathogenesis such as IL-9, IL-27, IL-15 and IL-2 (Figure 1D). Altogether, the cytokine *milieu* in RA indicated a highly inflammatory environment crucial for sustaining and exacerbating RA pathology.

**Figure 1.**
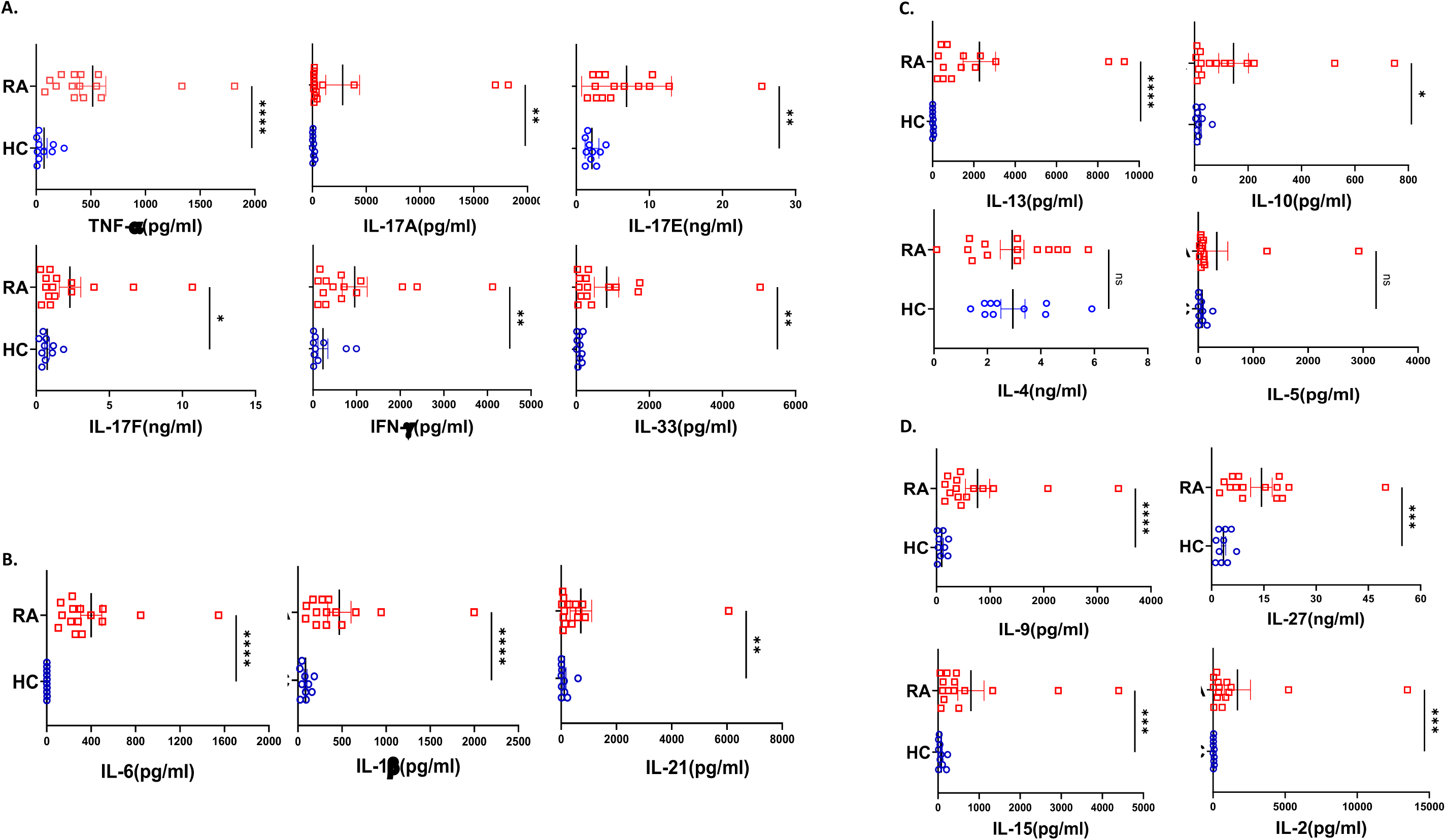
Cytokine analyses in RA patients and healthy controls. Representative figures from 20 T cell cytokines were analyzed in RA patients’ plasma (n=15) and healthy controls (n=10) and are represented as graphical plots. Inflammatory cytokines including TNF-α, IL- 17A, IL-17E, IL-17F, IFN-γ and IL-33 were significantly elevated in RA patients as compared to HC (A). In addition, Th17 driving cytokines such as IL-6, IL-1β and IL-21 were also significantly higher in RA patients with respect to healthy controls (B). Amongst the anti- inflammatory cytokines, only IL-13 and IL-10 was elevated in RA patients while IL-4 and IL- 5 did not show any difference between the two groups (C). RA patients also displayed higher expression of cytokines playing diverse roles in regulating and maintaining RA pathogenesis such as IL-9, IL-27, IL-15 and IL-2 in contrast to healthy controls (D). Mann-Whitney U Test was performed to compare between the two groups, p<0.05 was considered statistically significant (*), p<0.01 was considered to be very significant (**), p<0.001 was considered to highly significant (***), p<0.0001 was considered extremely significant (****) ns, not significant. Error bar represents SEM.

### RA CD4^+^ T cells are major inflammatory cytokine expressers

Considering the seminal status of CD4^+^ T cells in mediating RA-associated inflammation, we examined for the intracellular expression of various cytokines in these cells. Upon activation, RA CD4^+^ T cells derived from PBMCs expressed significantly higher inflammatory cytokines such as IL-17, TNF-α and IFN-γ as compared to the healthy controls (Figure 2A-D). Additionally, we found significantly higher GMCSF levels, which is in line with previous reports and attributes to its crucial role in driving a potent inflammatory response in RA pathology (Figure 2G, H). Along with the inflammatory cytokines, we found significantly elevated levels of IL-17A, an effector molecule of Th17 cells, primal in mediating RA pathology (Figure 2E, F). Interestingly, most of the RA CD4^+^ T cells were co-expressing the inflammatory cytokines, suggesting a highly inflammatory phenotype and aberrant status (Figure 2K, L). In addition, higher expression of inflammatory cytokines in RA CD4^+^ T cells was in corroboration with our multiplex analysis of plasma cytokines. However, IL-10 expression in RA CD4^+^ T cells was insignificant as compared to healthy controls, again suggesting that these cells are incompetent in mounting an anti-inflammatory response to counter inflammation. Our data from the infiltrated CD4^+^ T cells in RA synovial fluid did not find any significant expression of inflammatory or anti-inflammatory cytokines, probably owing to effusion of the above cytokines. Taken together, our data indicated a covert ongoing systemic inflammation in RA patients, with CD4^+^ T cells specifically the Th17 phenotype playing a critical role.

**Figure 2.**
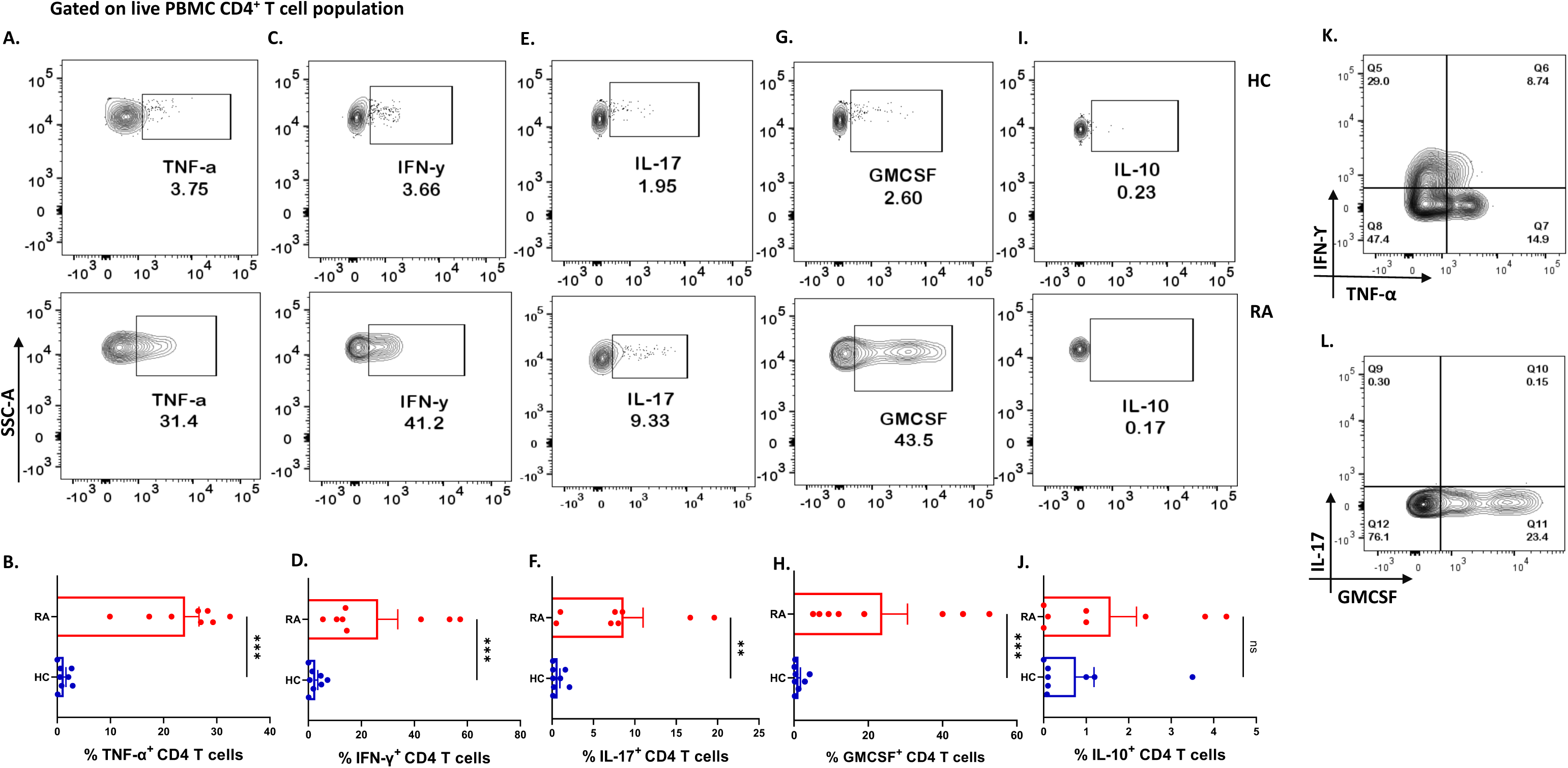
Differential expression of inflammatory and anti-inflammatory cytokines in RA CD4^+^ T cells. Intracellular staining of CD4^+^ T cells derived from RA PBMCs (n=8) displayed higher inflammatory cytokine expression than healthy controls (n=8). Representative flow cytometry plots show significantly elevated frequency of PBMC-derived CD4^+^ T cells expressing inflammatory cytokines such as TNF-α, IFN-γ, IL-17 and GMCSF (A, C, E, G & I) and cumulative graphical representation (B, D, F, H & J). Representative flow cytometry plots show non-significant expression of IL-10 in healthy controls and RA patients (I) and cumulative graphical representation (J). RA CD4^+^ T cell population co-express multiple inflammatory cytokines including, TNF-α, IFN-γ and GMCSF (K & L) where GMCSF^+^ and IL-17^+^ cells are gated on IFN-γ-TNF-α^+^ population. Mann-Whitney U Test was performed to compare between the two groups, p<0.05 was considered statistically significant (*), p<0.01 was considered to be very significant (**), p<0.001 was considered to highly significant (***), p<0.0001 was considered extremely significant (****) ns, not significant. Error bar represents SEM.

### RA CD4^+^ T cells mediate augmented osteoclastogenesis with higher RANKL expression

Along with inflammation, bone erosion is another key RA characteristic, primarily dependent on the augmented proliferation, differentiation and maturation of osteoclasts (the process of osteoclastogenesis), and is regulated by the RANK-RANKL pathway (Boyce and Xing 2008, Schett and Gravallese 2012). Apart from osteoblasts, previous studies suggest T cells express RANKL for normal physiological function (Anderson, Maraskovsky et al. 1997) and thus we wanted to compare its differential expression between CD4^+^ T cells of RA and healthy controls. Here, we found a significant increase in the expression of membrane-bound RANKL (mRANKL) in RA CD4^+^ T of PBMCs as compared to that of healthy controls (Figure 3A, B). Although RANKL expression was higher in the infiltrated CD4^+^ T cells of SFMCs (synovial fluid mononuclear cells) as compared to PBMC-derived CD4^+^ T cells of healthy controls, the expression levels were comparable between the PBMC and SFMC-derived CD4^+^ T cells (Figure 3A, C). Subsequently, we also confirmed the expression of membrane-bound RANKL on RA PBMC derived CD4^+^ T cells via confocal microscopy (Figure 3D). Altogether, our RANKL studies essentially indicated higher RANKL expression on RA CD4^+^ T cells, in turn suggesting higher osteoclastogenesis or bone erosion.

**Figure 3.**
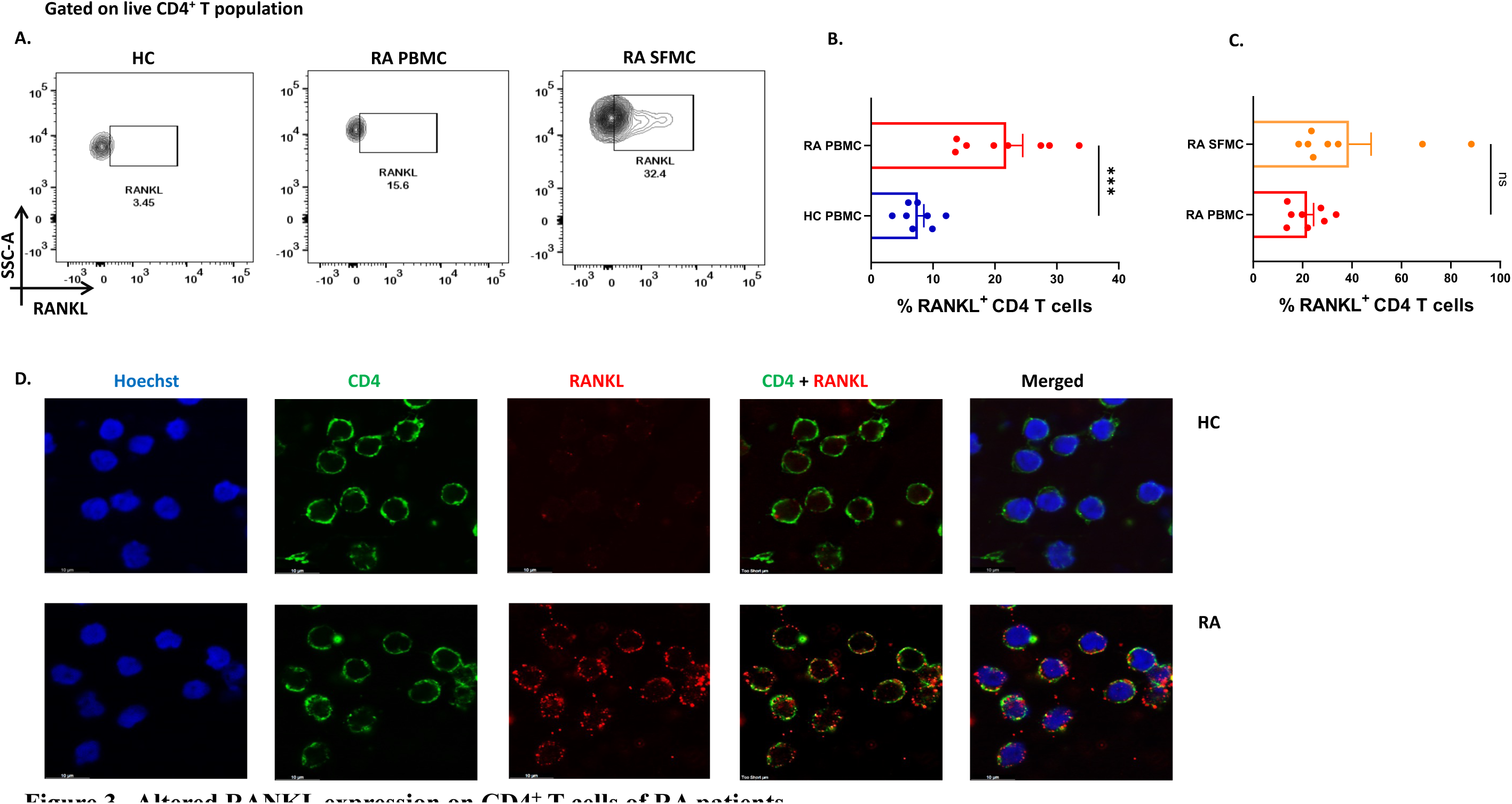
Altered RANKL expression on CD4^+^ T cells of RA patients. Surface staining of RANKL on CD4^+^ T cells of RA patients (n=8) showed higher expression than healthy controls (n=8). Representative flow cytometry plots show significantly elevated frequency of RANKL^+^ cells derived from CD4^+^ T cells of both peripheral blood and synovial fluid of RA patients (A). Cumulative graphical representation comparing RANKL^+^ CD4^+^ T cells of RA and HC PBMC (B) and between CD4^+^ T cells derived from RA PBMC and RA SFMC (C). Images acquired in TCS SP5 Leica confocal microscope show higher expression of membrane RANKL (mRANKL) on the surface of CD4^+^ T cells of RA patients as compared to healthy controls (D). Mann-Whitney U Test was performed to compare between the two groups, p<0.05 was considered statistically significant (*), p<0.01 was considered to be very significant (**), p<0.001 was considered to highly significant (***), p<0.0001 was considered extremely significant (****) ns, not significant. Error bar represents SEM.

### IL-21 and/or IL-23 modulates inflammatory response in RA CD4^+^ T cells

So far, our understanding of RA pathology involved characterization of two key attributes of RA CD4^+^ T cells i.e. higher inflammation and the capability of extensive bone resorption. Our results strongly indicated a dominant Th17 phenotype expressing IL-17, TNF-α, GM-CSF, IFN-γ etc. amongst inflammatory cytokines and RANKL predominantly in the IL-17 subset and FasL in both the IL-17 and IFN-γ compartment (data not shown). Subsequently, we questioned the involvement of specific driving and stabilizing cytokine(s) and associated pathways in modulating both the processes by examining the augmented levels of these cytokines in the RA plasma. Here, we examined and found significantly higher expression of cytokines, IL-21 and IL-23 (both p19 and p40 subunits) in RA CD4^+^ T cells as opposed to healthy controls (Figure 4A-F). Consequently, we selectively inhibited the cytokines IL-21 and or IL-23 using neutralization antibodies during activation of RA CD4^+^ T cells and analyzed for altered expression of inflammatory cytokines. Interestingly, we found significant reduction in both IL-17 and TNF-α expression with both αIL-21 and αIL-23 treatment, independently as well as in combination (Figure 5A-H). The significant inhibition in the signature cytokine IL-17 and the pleiotropic cytokine TNF-α strongly suggested that cytokines IL-21 and IL-23 work in tandem to drive a sustained inflammatory response. With respect to IFN-γ expression, we found significant decrease with αIL-21 treatment and in combination only (Figure 5I-L). RA CD4^+^ T cells did not display any alteration in GMCSF expression with inhibition of any of the cytokines. The above findings strongly suggested that the IL-21/23 axis plays a major role in contributing to the exaggerated inflammation of RA CD4^+^ T cells. Significantly, its inhibition resulted in reducing the signature and pleiotropic cytokine IL-17 and TNF-α, had selective effect on the Th17 cell inhibitory cytokine IFN-γ, while having no effect on the adjuvant cytokine GM-CSF.

**Figure 4.**
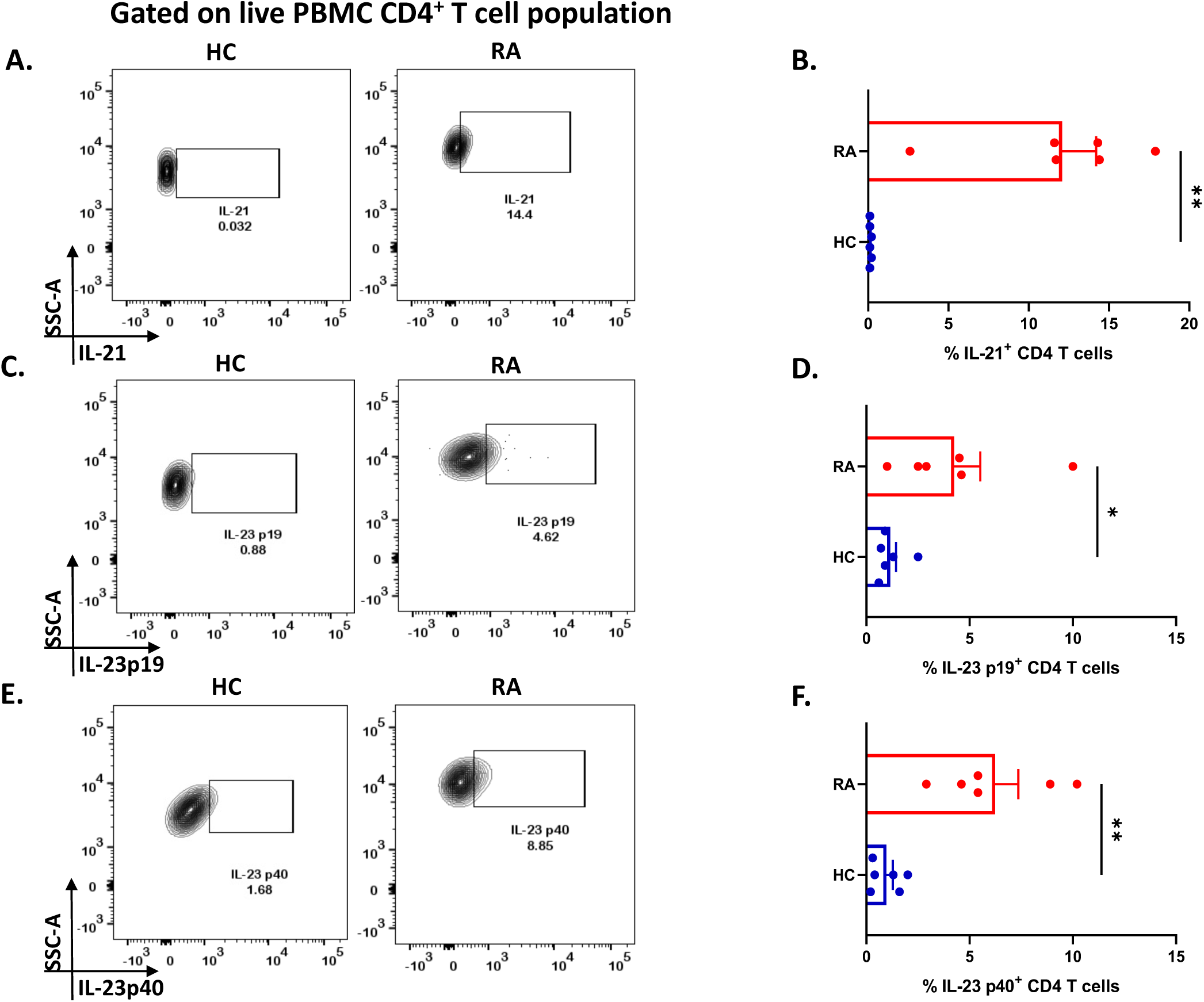
IL-21 and IL-23 expression in RA CD4^+^ T cells. Intracellular staining of inflammatory cytokines, IL-21 and both subunits of IL-23 displayed significantly higher expression in RA CD4^+^ T cells (n=6) upon PMA/Ionomycin stimulation as compared to healthy controls (n=6). Representative flow cytometry plots show significant elevation in IL-21, IL- 23p19 and IL-23p40 expression in CD4^+^ T cells derived from RA and HC PBMCs (A, C and E) and found statistically significant in the corresponding graphical represenatation (B, D and F). Mann-Whitney U Test was performed to compare between the two groups, p<0.05 was considered statistically significant (*), p<0.01 was considered to be very significant (**), p<0.001 was considered to highly significant (***), p<0.0001 was considered extremely significant (****) ns, not significant. Error bar represents SEM.

**Figure 5.**
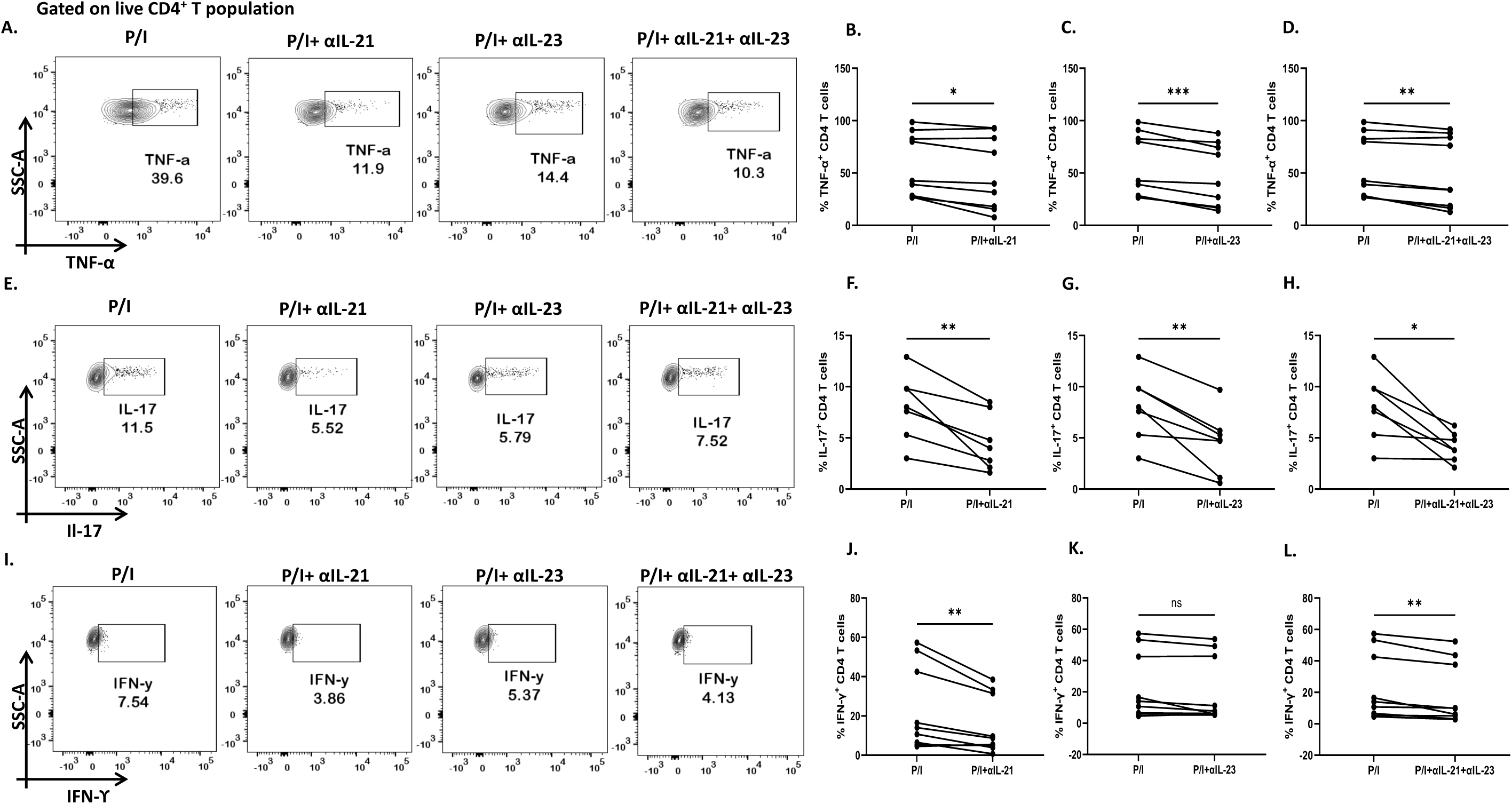
Modulation of inflammatory cytokines in RA CD4^+^ T cells by αIL-21 and αIL-23. Representative figures showing modulation of inflammatory cytokines in RA PBMC derived CD4^+^ T cells (n=8) with αIL-21 and αIL-23 inhibition along with PMA/Ionomycin stimulation. Stimulated cells selectively inhibited with αIL-21 show significant decrease of cytokines, TNF-α, IL-17 and IFN-γ with respect to only PMA/Ionomycin treated as depicted in the representative flow cytometry plots (A, E and I) and in cumulative graphical representation (B, F and J). Significant reduction of TNF-α and IL-17 expression was also observed with αIL-23 inhibition in RA CD4+ T cells as represented in flow cytometry plots (A and E) and cumulative graphs (C and G). IFN-γ expression did not show significant difference with αIL-23 treatment (I and K). Expression of TNF-α, IL-17 and IFN-γ reduced significantly with combined treatment of αIL-21 and αIL-23 as depicted in the representative flow cytometry plots (A, E and I) along with cumulative graphical representation (D, H and L). Paired T Test was performed to compare between the two groups, p<0.05 was considered statistically significant (*), p<0.01 was considered to be very significant (**), p<0.001 was considered to highly significant (***), p<0.0001 was considered extremely significant (****) ns, not significant. Error bar represents SEM.

### IL-21/IL-23 axis regulates RANKL expression on RA CD4^+^ T cells

As our findings suggested that inflammation was regulated by the cytokines, IL-21 and IL-23, we furthered our hypothesis to examine if these cytokines could also be responsible for upregulating membrane-bound RANKL expression. To that end, we inhibited IL-21 and IL- 23 (p19 and p40) using neutralization antibodies during activation. Surprisingly, we found that inhibition of IL-21 and IL-23 also reduced RANKL expression significantly on RA CD4^+^ T cells derived from PBMCs and SF, both independently and when inhibited together (Figure 6A-H). Taken together, this suggested that IL-21 and IL-23 were responsible for modulation of inflammatory cytokines, IL-17, TNF-α, IFN-γ as well as RANKL, thus establishing and validating our hypothesis; IL-21/IL-23 axis dictates the pathways regulating inflammation and RANKL expression in aberrant CD4^+^ T cells in RA.

**Figure 6.**
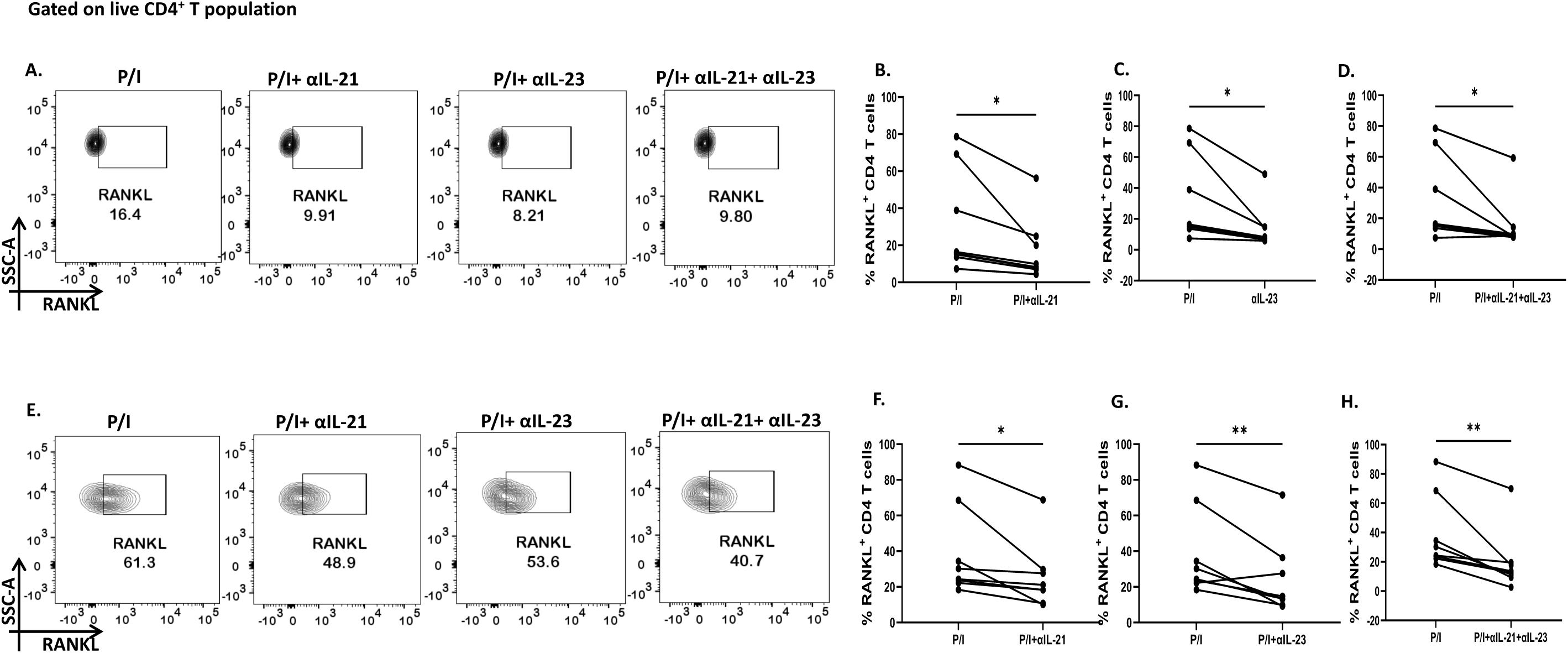
RANKL modulation with αIL-21 and αIL-23 treatment. Representative figures showing modulation of RANKL expression in RA CD4^+^ T cells (n=8) derived from PBMCs and SFMCs with αIL-21 and αIL-23 treatment along with PMA/Ionomycin stimulation. Both PBMC and SFMC derived CD4^+^ T cells displayed significant downregulation of RANKL with αIL-21 treatment as shown (A and E) and in graphical plots (B and F). In addition, RANKL expression decreased significantly in PBMC and SFMC derived CD4^+^ T cells with αIL-23 treatment and with αIL-21+ αIL-23 in flow cytometry plots (A and E) and graphical plots (C, D, G and H). Paired T Test was performed to compare between the two groups, p<0.05 was considered statistically significant (*), p<0.01 was considered to be very significant (**), p<0.001 was considered to highly significant (***), p<0.0001 was considered extremely significant (****) ns, not significant. Error bar represents SEM.

### IL-21 & IL-23 modulates IL-17 and RANKL in e*x vivo* polarised human Th17

We wanted to confirm that IL-21 and IL-23 are indeed responsible for modulating the inflammatory cytokine response and RANKL expression and that our findings could have clinical co relates and significance. Thus, we furthered our findings in *ex vivo* differentiated human Th17 cells, selecting this subtype on the basis of its long known role in RA pathogenesis. Towards this, we first differentiated human Th17 cell and characterized it based on the expression of its signature cytokine IL-17, master transcription factor Rorγt and surface marker CCR6 (Figure 7A, C and I). We found statistical similarity between all the tested batches of the same (Figure 7B, D). Subsequently, we analyzed for IL-17 expression with αIL- 21 and αIL-23 inhibition and found a significant reduction in these cells (Figure 7E, F). These results were in line with our findings from patient CD4^+^ population. In addition, the *ex vivo* differentiated Th17 cells co-expressed RANKL on its surface, along with CCR6 (Figure 7I, J) and its expression decreased significantly upon inhibition of both, IL-21 and IL-23 (Figure 7I- L). These results implicitly and explicitly established the crucial role played by the IL-21/23 axis in both inflammation and RANKL expression in both terminally differentiated physiological Th17 and patient derived pathological derived Th17 cells. Altogether, our *ex vivo* studies reinforced our hypothesis about the crucial role played by IL-21 and IL-23 in exaggerating the inflammatory response as well as RANKL expression in Th17 cells in physiology and RA.

**Figure 7.**
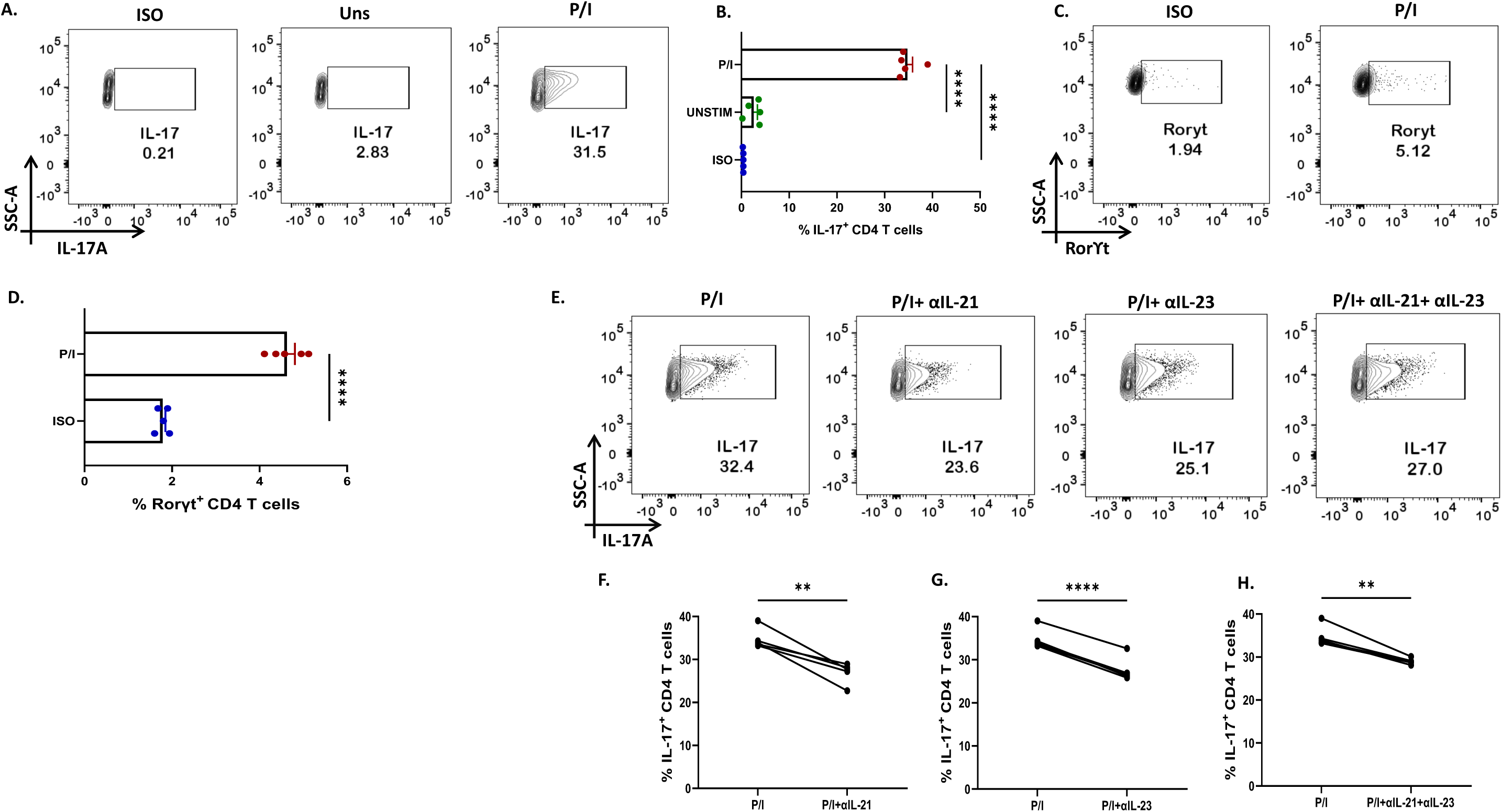
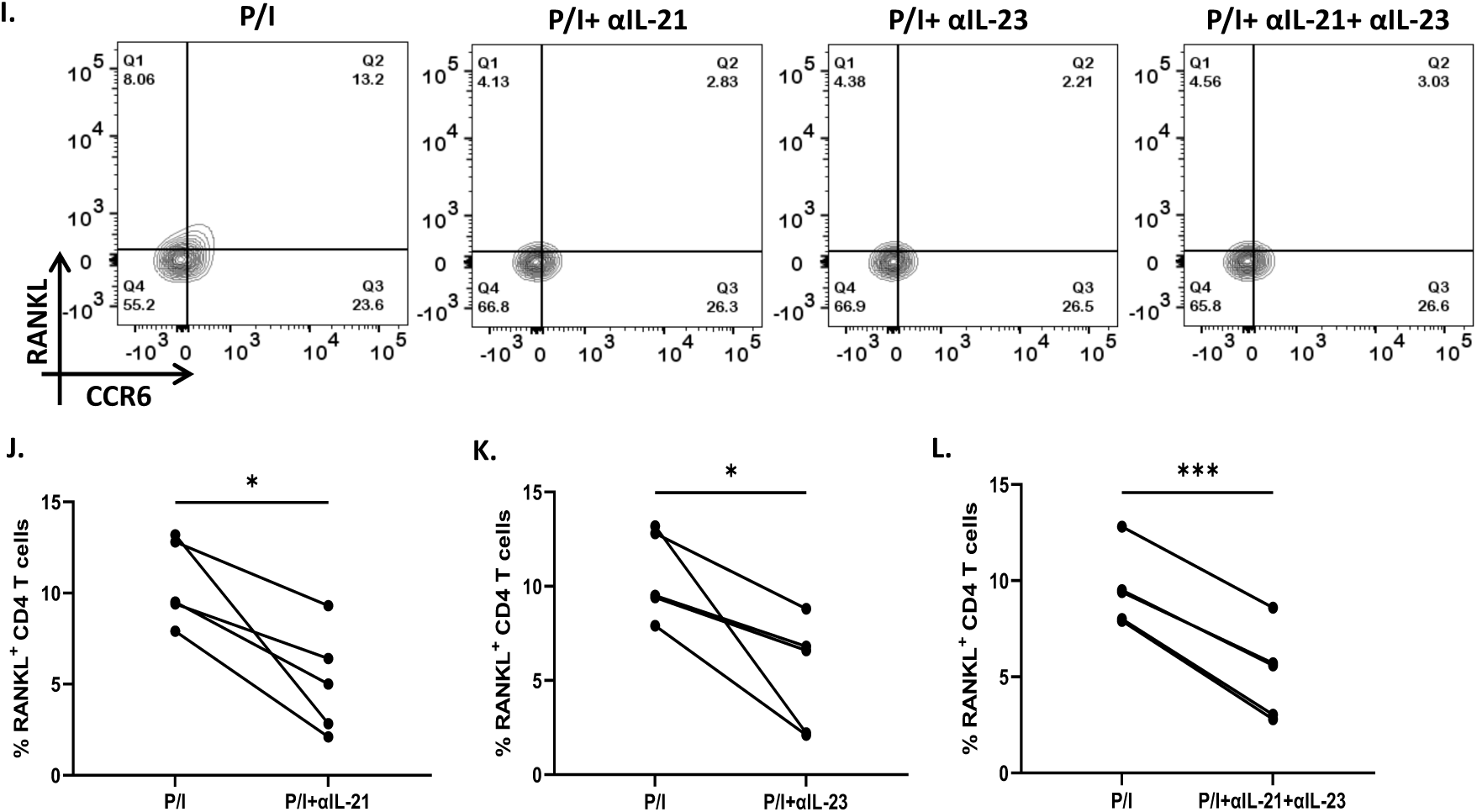
IL-21 and IL-23 modulate signature cytokine and RANKL expression of *ex vivo* polarized human Th17 cells. Negatively isolated CD4^+^ T cells derived from PBMCs were activated with αCD3/28 stimulation, polarizing cytokines and neutralizing antibodies for 10 days, characterized for Th17 phenotype and examined for altered expression of cytokines and RANKL with αIL-21 and αIL-23 treatment upon PMA/Ionomycin stimulation. Post 10-day polarization, CD4^+^ T cell population showed significant expression of IL-17, signature cytokine for Th17 subset as compared to the unstimulated cells represented in flow cytometry plots (A) as well as the graphical plots (B). In addition, our representative figures display significant expression of Th17 phenotype’s master transcription factor, RorγT (C and D). Mann-Whitney U Test was performed to compare between the two groups. Treatment of these cells with both, αIL-21 and αIL-23 showed significant decrease in IL-17 expression, independently as well as in combination as is represented in flow cytometry plots (E) and graphical plots (F, G and H). In addition, RANKL expression on CCR6^+^ cells also decreased significantly with both αIL-21 and αIL-23 treatment as represented in flow cytometry plots (I) and graphical plots (J, K and L). Paired T Test was performed to compare between the two groups, p<0.05 was considered statistically significant (*), p<0.01 was considered to be very significant (**), p<0.001 was considered to highly significant (***), p<0.0001 was considered extremely significant (****) ns, not significant. Error bar represents SEM.

### IL-21/23 axis modulates inflammatory cytokines and RANKL via PI3K/p-Akt pathway

Based on our above findings, we hypothesized that both IL-21 and IL-23 regulated the inflammatory response and RANKL expression in RA CD4^+^ T cells via common downstream pathway. Previous reports suggest PI3K/p-Akt pathway to be common and critical for both the cytokines and thus we next examined for the altered expression of pAkt-1 with αIL-21 and αIL-23 inhibition. Interestingly, we found significant decrease in p-Akt1 expression with both αIL-21 and αIL-23 treatment, independently as well as when added together, thus indicating towards p-Akt1 being a key signalling molecule modulated by the cytokines, IL-21 and IL-23 (Figure 8A-D). Subsequently, we inhibited p-Akt1 expression with pAkt1/2 kinase inhibitor during TCR stimulation and examined for altered inflammatory cytokine and RANKL expression. Not surprisingly, there was significant reduction of inflammatory cytokines such as TNF-α, IL-17 and IFN-γ in RA CD4^+^ T cells with p-Akt1 inhibition, thus confirming our hypothesis (Figure 8E, G and I). In addition, we also found significant decrease in RANKL expression of RA CD4^+^ T cells with p-Akt1 inhibition (Figure 8K) and report statistical significance in all the tested batches (Figure 8F, H, I and J). We further validated our hypothesis through confocal imaging where inhibition of Akt1 phosphorylation restricted nuclear translocation (Figure 8M). Although we understand that the expression of proteins are regulated by multiple signalling pathways, our findings above primarily dissected one of the critical pathways employed by IL-21/23 axis to regulate two critical processes in RA CD4^+^ T cells i.e. hyper inflammatory cytokines and augmented RANKL expression.

**Figure 8.**
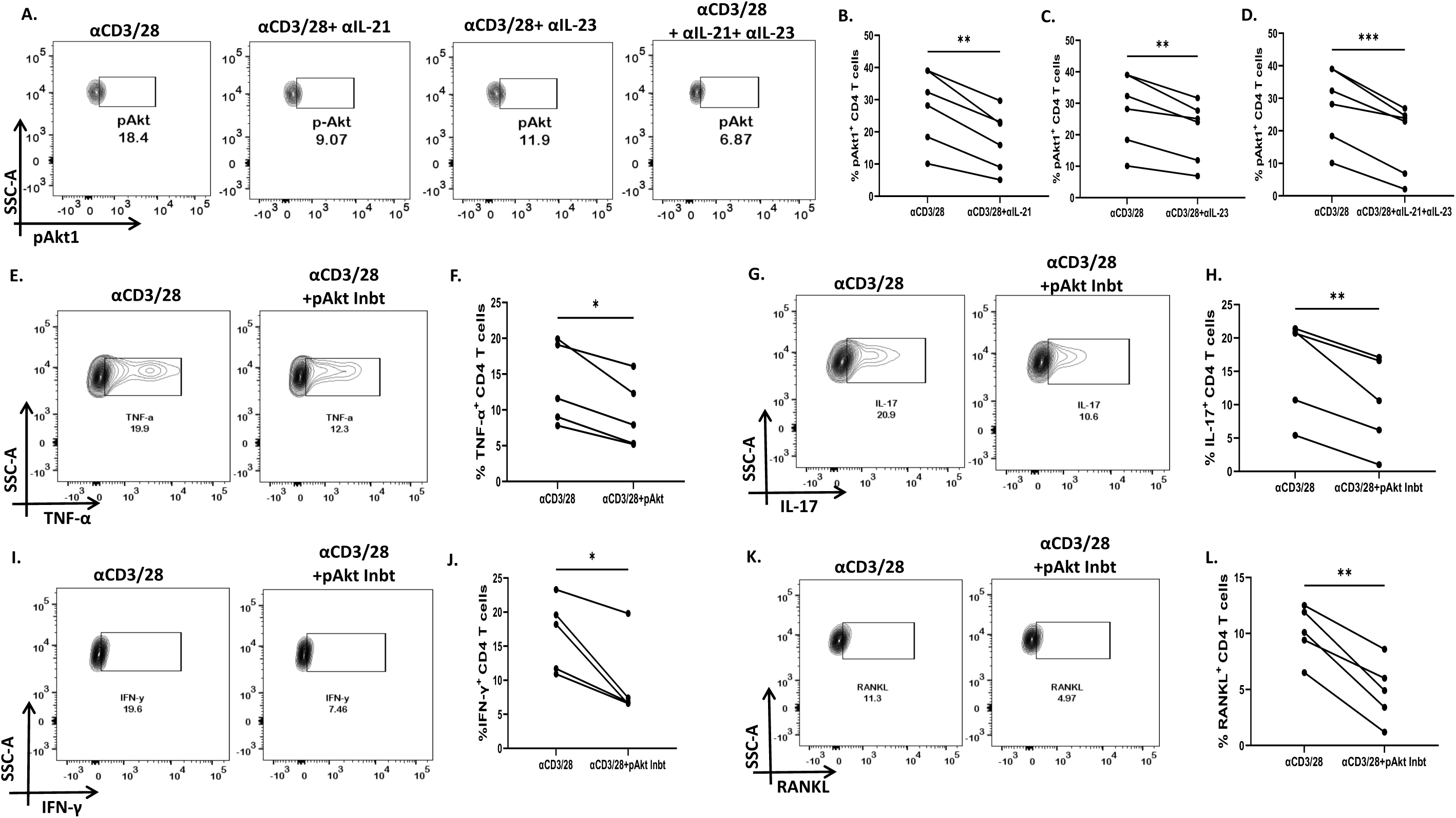
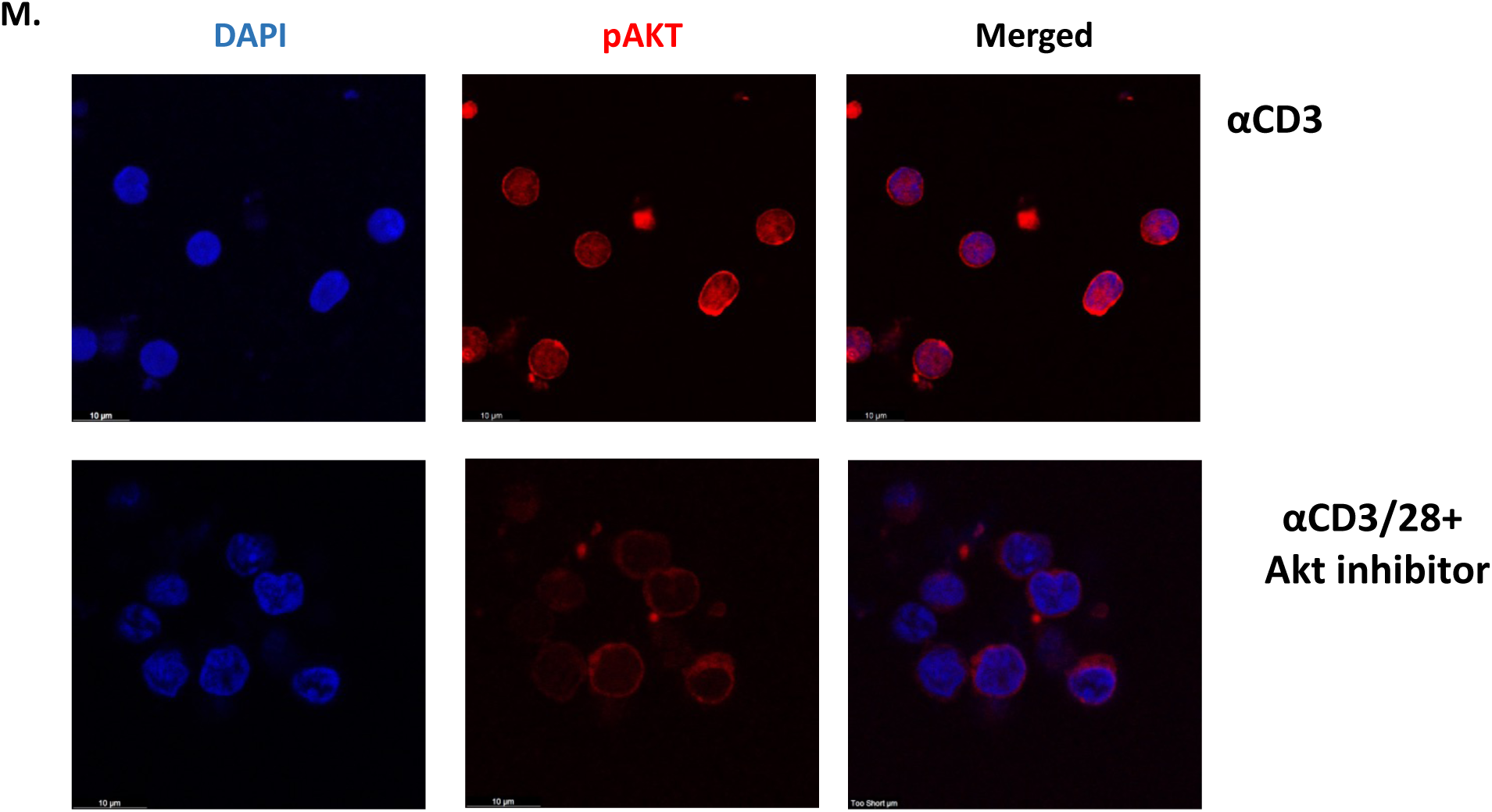
IL-21 and IL-23 modulates inflammatory cytokines and RANKL expression via pAkt1. RA CD4^+^ T cells were stimulated with αCD3/28 along with αIL-21 and αIL-23 treatment for 24 hours and examined for alteration in pAkt1 expression and later, analyzed for altered expression of cytokines and RANKL with addition of Akt1/2 kinase inhibitor. Representative figures showing significant decrease in pAkt1 expression with αIL-21 and αIL- 23 treatment, independently as well as in combination as shown in flow cytometry plots (A) and graphical plots (B, C and D). Addition of 5µM pAkt1/2 inhibitor to these cells during αCD3 stimulation displayed significant decrease in TNF-α, IFN-γ, IL-17 and RANKL expression as represented in flow cytometry plots (E, G, I and K) and graphical plots (F, H, J and L). Paired T Test was performed to compare between the two groups, p<0.05 was considered statistically significant (*), p<0.01 was considered to be very significant (**), p<0.001 was considered to highly significant (***), p<0.0001 was considered extremely significant (****) ns, not significant. Error bar represents SEM.

## Discussion

Amongst an array of immune cells governing the RA microenvironment, CD4^+^ T cells have long been known to play a central role in its pathogenesis (Klarenbeek, de Hair et al. 2012, Chemin, Gerstner et al. 2019). Over a decade long or even higher course of disease progression, CD4^+^ T cells undergo various stages of dysfunctionality and mis-regulation of associated pathways, eventually leading to hyper-inflammation and extensive bone degradation. Our key objective in this study was to understand and establish the seminal role played by specific cytokines in regulating both, tissue inflammation and destruction with respect to CD4^+^ T cells.

Our preliminary studies essentially characterized the cytokine *milieu* in RA patients and here we report significantly elevated levels of multiple inflammatory cytokines as opposed to that of healthy controls. In addition, we found significantly higher levels of inflammatory cytokines specifically responsible for driving aberrant CD4^+^ T cell phenotypes along with other pleiotropic cytokines regulating and maintaining various aspects of RA pathology. This was further established by our findings of RA CD4^+^ T cells driving an exaggerated inflammatory response and also suggested that these cells are one of the key sources of the inflammatory cytokines. Although the RA cytokine *milieu* was essentially inflammatory, our multiplex analysis also indicated towards augmented levels of certain anti-inflammatory cytokines such as IL-13 and IL-10. However, as these cytokines could not lessen the exaggerated inflammation in RA, we assumed this to be a consequence of inadequate levels of anti- inflammatory cytokines or the highly unresponsive aberrant RA immune cells. Interestingly, most of the inflammatory cytokines such as TNF-α, IFN-γ and GMCSF were co-expressed by the CD4^+^ T cell population, strongly suggesting their aberrant status. IL-17 was found to be an exception in most cases with low but significant expression in RA patients as compared to healthy controls. In addition, our studies assessing cytokine levels in RA synovial fluid and the infiltrated CD4^+^ T cells in SFMCs did not find any significant expression of cytokines, possibly due their effusion into peripheral circulation.

Along with inflammation, extensive bone degradation is another characteristic associated with RA pathogenesis (Iwamoto and Kawakami 2022). As bone degradation or erosion is essentially governed by the imbalance between populations of osteoblasts and osteoclasts, the process of osteoclastogenesis is of critical importance in RA. In physiology, this process is mediated via RANK-RANKL interaction, where RANK is a transmembrane protein of the TNF family of receptors expressed on osteoclasts, RANKL being its ligand, primarily expressed on osteoblasts (Boyce 2013, Ono, Hayashi et al. 2020). Interestingly, RANKL is also expressed in both, soluble and membrane bound form in T cells (Anderson, Maraskovsky et al. 1997, Wong, Josien et al. 1997). Thus, we next questioned the capability of RA CD4^+^ T cells in expressing membrane bound RANKL and not surprisingly, found significantly higher expression as opposed to that of CD4^+^ T cells from healthy controls. When compared between the RANKL expression on infiltrated CD4^+^ T cells of synovial fluid and peripheral blood, we did not find any significant difference, possibly suggesting that these cells have effused out from the target site into circulation. RANKL expression was evenly distributed on the surface of RA CD4^+^ T cells, but were not visible on CD4^+^ T cells of healthy controls.

Till here, we had a distinct impression of an aberrant RA CD4^+^ T cell; it was highly inflammatory and expressed significantly higher RANKL. Subsequently, we questioned the possibility of a common cytokine signalling pathway responsible for amplifying both, inflammatory cytokines and RANKL expression. With the known involvement of Th17 cells in precipitating RA (van den Berg and McInnes 2013, van Hamburg and Tas 2018), we primarily based our assumption on Th17-polarising cytokines playing a critical role. Interestingly, neutralization of both, IL-21 and IL-23 in RA CD4^+^ T cells not only yielded significant reduction in the expression of inflammatory cytokines TNF-α, IL-17 and IFN-γ, but also downregulated RANKL expression, directly indicating towards the critical role played by these cytokines in augmenting the inflammatory response as well as osteoclastic activity. Here, we did not find any significant change in the expression of GMCSF with IL-21 and/or IL-23 neutralization, thus suggesting an alternative pathway regulating its expression.

IL-21/23 axis regulates various downstream molecules via multiple pathways (Cho, Kang et al. 2006, Leonard and Wan 2016, Pastor-Fernandez, Mariblanca et al. 2020). Thus, we aimed to identify the common downstream signalling pathway regulating the expression of both, inflammatory cytokines and RANKL. We found low expression of pSTAT3 in RA CD4^+^ T cells (Supplementary figure 6), thus negating its role in regulating the expression of inflammatory cytokines and RANKL. However, the expression of p-Akt1 reduced significantly with IL-21 and IL-23 inhibition, suggesting PI3K/Akt pathway playing a central role in modulating inflammatory cytokine and RANKL expression. Further inhibition of p- Akt1 expression with pAkt1/2 kinase inhibitor not only downregulated inflammatory cytokines but also resulted in decreased RANKL expression, thus confirming that IL-21/23 axis employs PI3K/p-Akt1 pathway for regulation of CD4^+^ T cell mediated inflammation and osteoclastogenesis in RA.

This study essentially addresses the modulatory role played by cytokines, IL-21 and IL- 23 in mediating two critical features associated with RA pathogenesis: inflammation and increased osteoclastogenic activity. Although we understand that this study is based on a very small sample size and should be validated further in a larger cohort for better understanding, we must emphasize on the therapeutic implications of this study. Altogether, IL-21/23 axis mediated via PI3K/p-Akt1 signalling pathway sheds light into one of the crucial pathways indispensable for the aberrant status of CD4^+^ T cells in RA.

## Ethics Statement

The study bearing reference number 76/HEC/18 was conducted according to the guidelines of the Declaration of Helsinki and approved by the Institutional Human Ethics Committee and Institutional Review Board (IRB) Institute of Life Sciences. Written consent informed to participate in this study was provided by the participants.

## Data Availability Statement

All the relevant clinical and laboratory data will be provided on request from researchers with Informed consent from patient.

## Author Contributions

Experimental Design and Conceptualization: SD, GB and SS; Sample Collection, Processing and Clinical Scoring: JRP, PP and PKB; Flow Cytometry Experiments: GB and RJ; Cytokine Multiplexing: GB, SS and SKS; Confocal experiment Design, Data Collection and analysis: SD, RJ and GB; *Ex vivo* human Th17 differentiation experiments: GB and GMJ

## Supporting information

Reagents used in the study

## Data Availability

All the relevant clinical and laboratory data will be provided on request from
researchers with Informed consent from patient.

## Acknowledgements

We would like to thank Dr. Subhasmita Mohanty for her assistance and valuable suggestions in flow cytometry and confocal imaging experiments. We would also like to thank Mr. Paritosh Nath, technical assistant in flow cytometry facility and Mr. Bhabani Sankar Sahoo, technical assistant in Confocal Imaging facility for their assistance.

## Figure legends

