## Supplementary Figures for "IL-21/23 axis modulates inflammatory cytokines and RANKL expression in RA CD4^+^ T cells via p-Akt signaling"

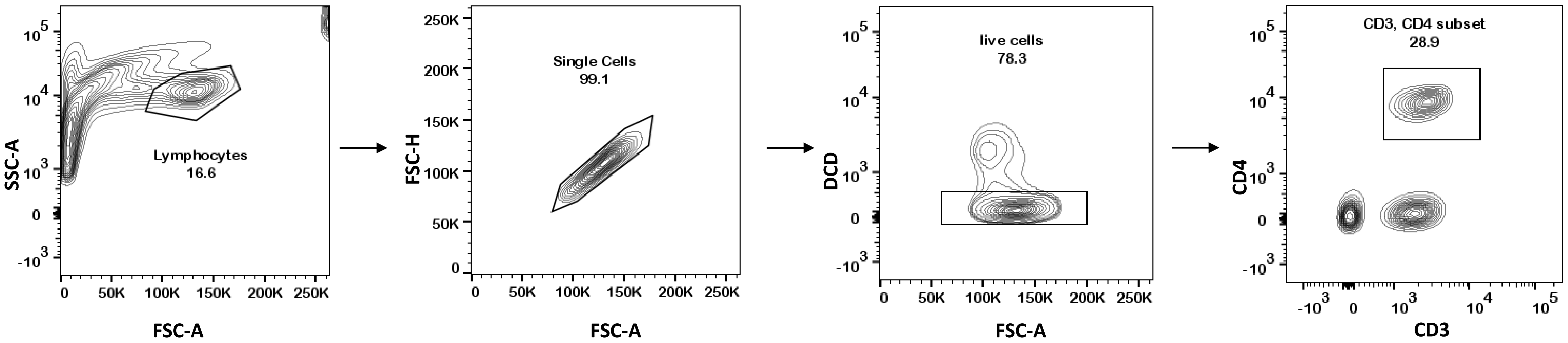


S1: Gating strategy for RA and HC PBMC derived $CD4^+$ T cells

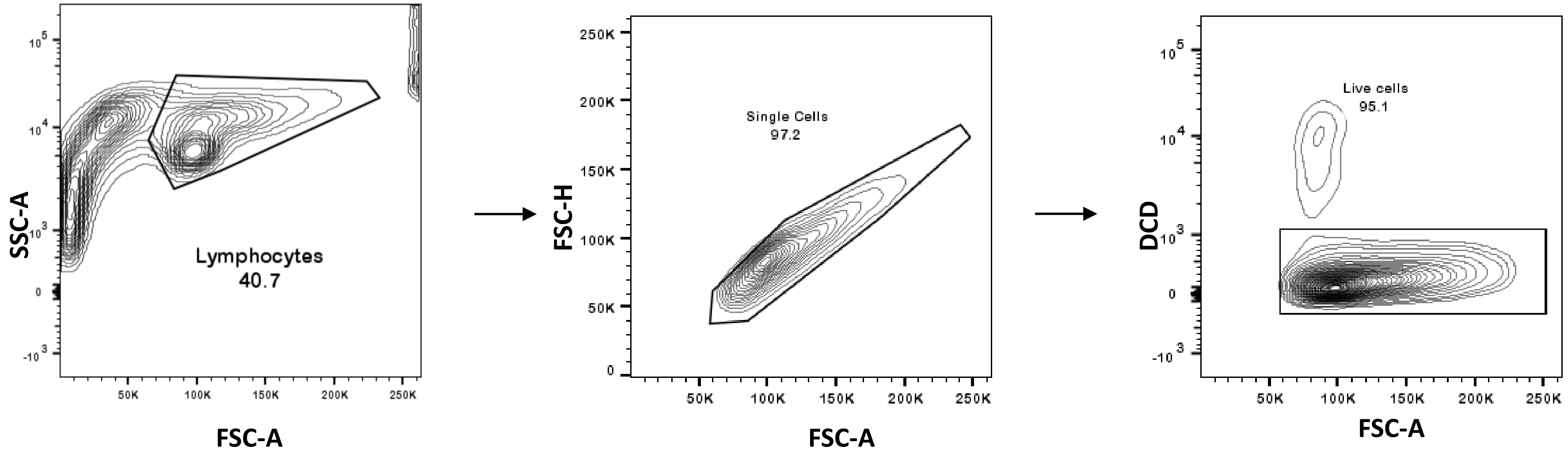


S2: Gating strategy for *ex vivo* differentiated CD4⁺ T cell experiments

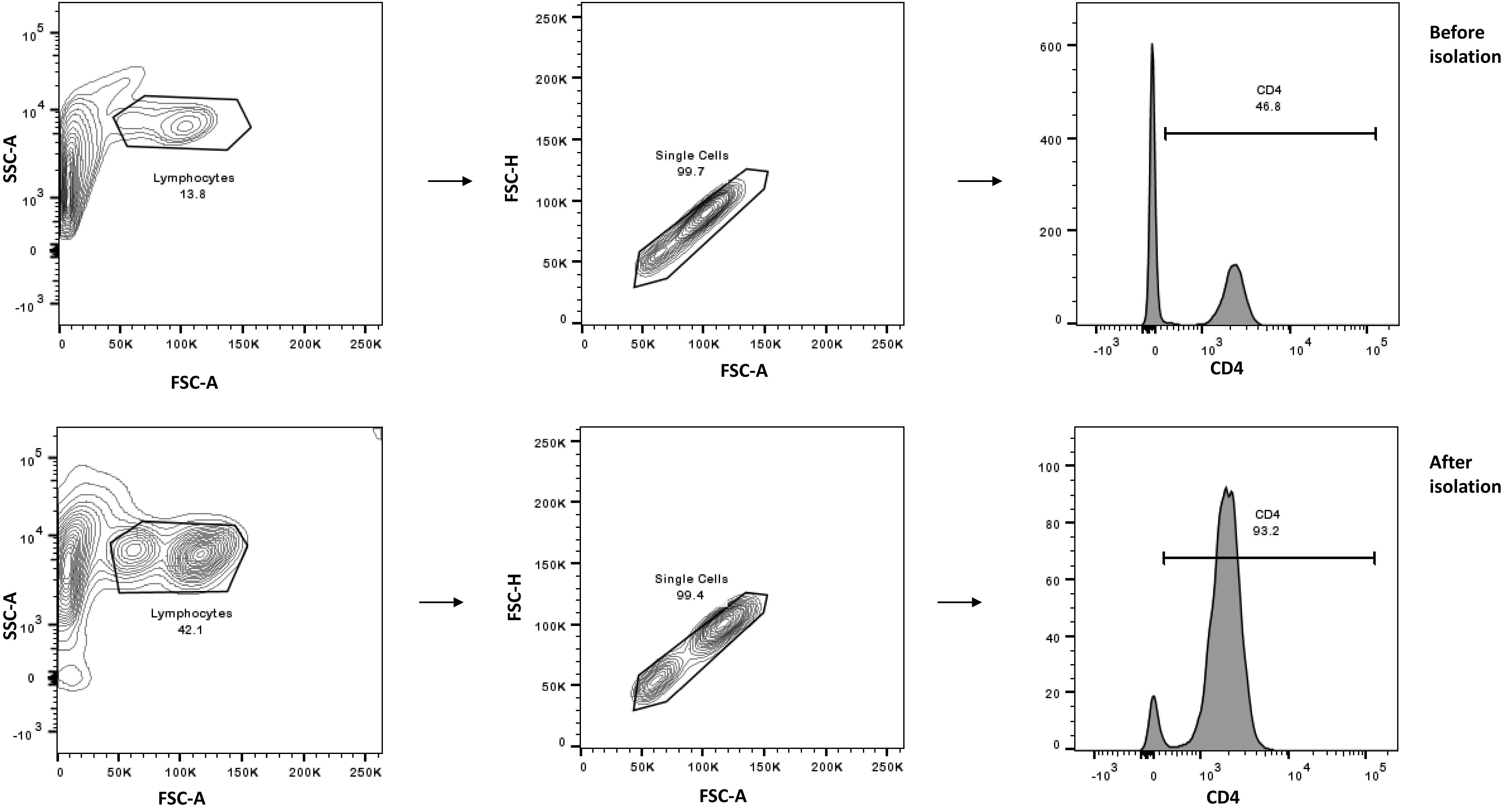


S3: CD4⁺ T cell purity validated for *ex vivo* experiments

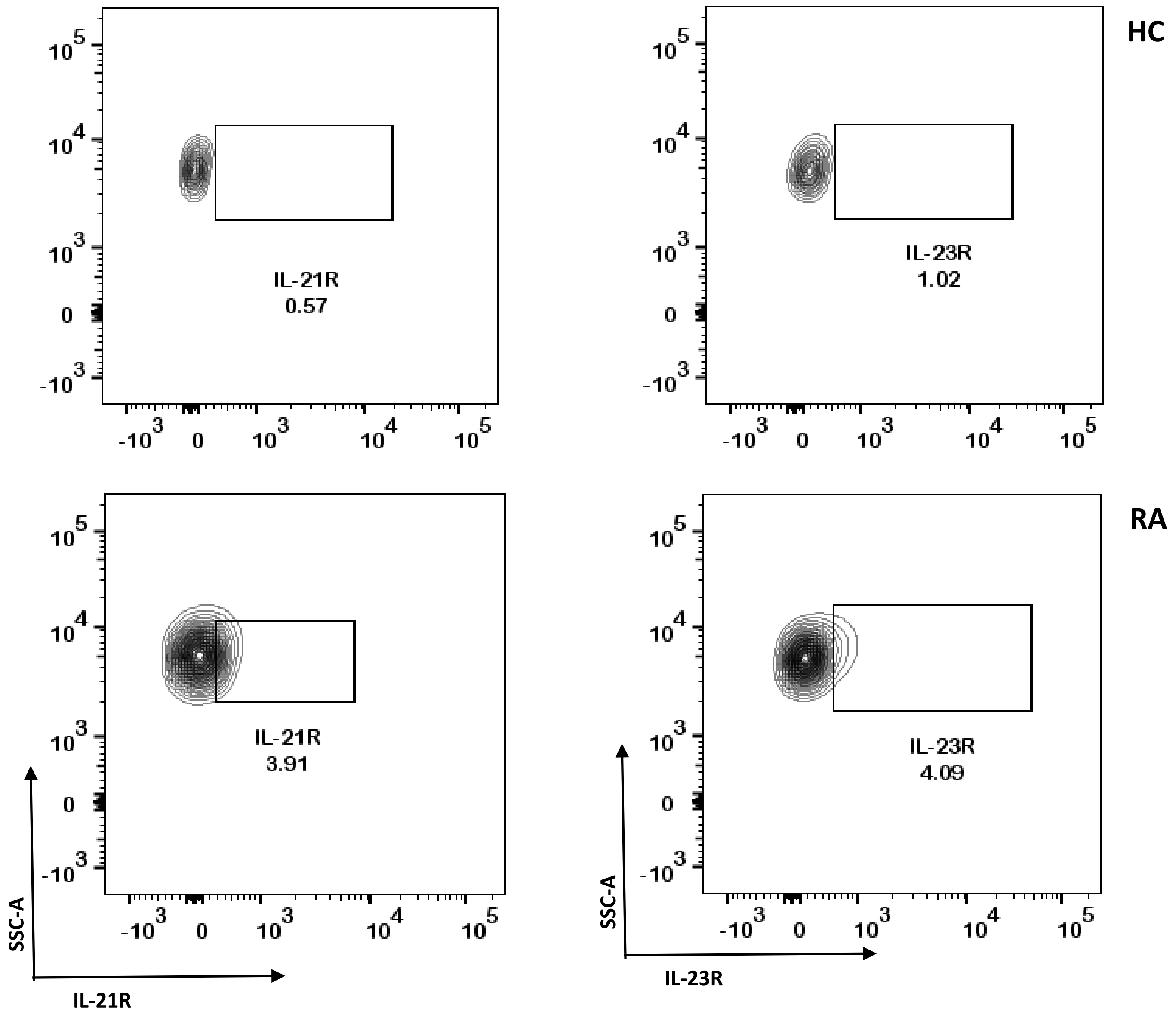


S4: Expression of IL-21R and IL-23R in HC and RA $CD4^+$ T cells

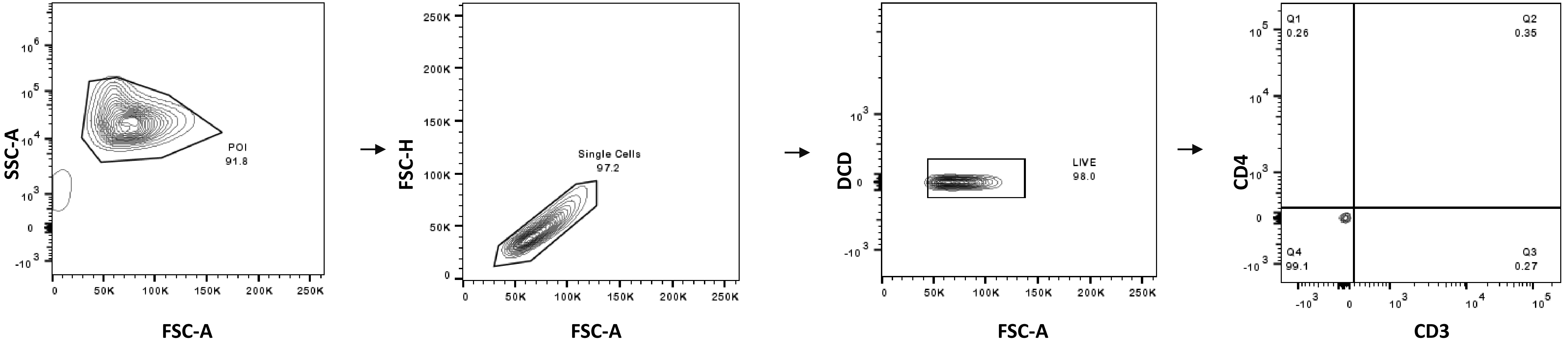


S5: Absence of T cell marker proteins in OA SF derived cells

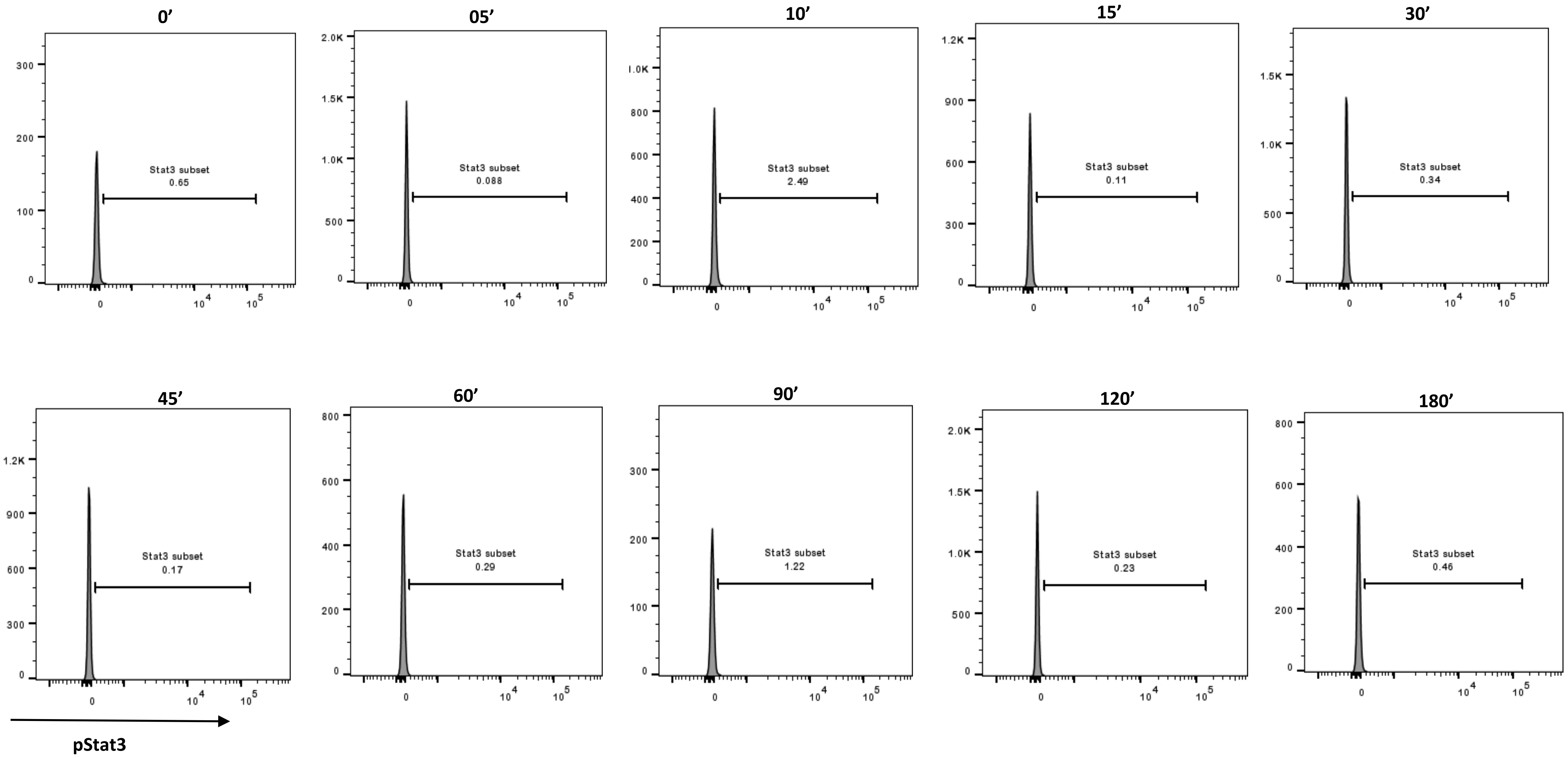


S6: Time kinetics of p-Stat3 expression in RA $CD4^+$ T cells
