## Supplementary material for "IL-21/23 axis modulates inflammatory cytokines and RANKL expression in RA CD4^+^ T cells via p-Akt signaling": Reagents used in the study

| Reagent | Catalogue No. |
|---|---|
| Milliplex MAP Human Th17 Magnetic Bead Panel-Immunology Multiplex Assay | Millipore HTH17MAG-14K |
| Dynabeads ™ Untouched Human CD4 T cells Kit | Invitrogen 11346D |
| BD Cytofix/Cytoperm™ | BD Biosciences 554714 |
| eBiosciences™ FoxP3/Transcription Factor Staining Buffer Set | Invitrogen 00-5523-00 |
| Poly-L-Lysine 0.01 | Sigma Aldrich P4707 |
| ProLong™ Gold Antifade Mountant with DAPI | Invitrogen P36935 |
| Hoechst Stain | Sigma Aldrich H6024 |
| Hyaluronidase from bovine testes | Sigma Aldrich H3506-100MG |
| RPMI 1640 | PAN-BIOTECH P04-16520 |
| Fetal Bovine Serum | PAN-BIOTECH P30-1402 |
| Zombie Violet™ Fixable Viability Kit | Biolegend 423113 |
| DPBS, w/0: Ca and Mg | PAN BIOTECH P04-36500 |
| Akt1/2 Kinase Inhibitor | Sigma Aldrich A6730-5MG |
| FlowJO Version 10.8 | BD Biosciences |
| Graph Pad Prism 9 | Dotmatics Pvt Ltd. |

| **Recombinant Human Cytokines** | **Catalogue No.** |
|---|---|
| IL-6 Protein Human Recombinant | Prospec Bio<br>CYT-213 |
| IL-1β Protein Human Recombinant | Prospec Bio<br>CYT-208 |
| TGF-β Protein Human Recombinant | Prospec Bio<br>CYT-716 |
| IL-21 Protein Human Recombinant | Prospec Bio<br>CYT-408 |
| IL-23 Protein Human Recombinant | Prospec Bio<br>CYT-050 |

| Fluorophore-tagged antibody | Catalogue No. Antibody | Isotype |
|---|---|---|
| IFN-γ BV480 | BD Biosciences 566100 | BD Biosciences 565652 |
| TNF-α PECF594 | BD Biosciences 562784 | BD Biosciences 562292 |
| IL-21R PECF594 | BD Biosciences 564122 | BD Biosciences 562292 |
| RANKL APC | Biolegend 347508 | Biolegend 400322 |
| IL-23R PE | R&D FAB14001P | R&D IC0041P |
| CD3 AF700 | Biolegend 317340 | Biolegend 400248 |
| IL-17 PerCPCy5.5 | BD Biosciences 560799 | BD Biosciences 552834 |
| IL-23p40 PE | Biolegend 501806 | R&D IC002P |
| RANKL PE | Biolegend 347503 | Biolegend 400311 |
| IL-23p19 PE | R&D IC17161P | R&D IC0041P |
| IL-21 AF647 | BD Biosciences 562043 | BD Biosciences 557714 |
| GMCSF AF647 | BD Biosciences 562257 | BD Biosciences 557906 |
| T-bet AF488 | BD Biosciences 561266 | Ebioscience 53-4714-42 |
| RorγT BV650 | BD Biosciences 563424 | BD Biosciences 563437 |
| IL-10 PE | BD Biosciences 559330 | BD Biosciences 559317 |
| CD4 PECy7 | Biolegend 300512 | Ebioscience 25-4714-42 |
| pAkt1 APC | Ebioscience 17-9715-42 | Ebioscience 17-4724-81 |
